# Tracking the mental health of home-carers during the first COVID-19 national lockdown: evidence from a nationally representative UK survey

**DOI:** 10.1101/2021.01.25.21250437

**Authors:** Elise Whitley, Kelly Reeve, Michaela Benzeval

**Affiliations:** MRC/CSO Social and Public Health Sciences Unit, University of Glasgow, Berkeley Square, 99 Berkeley Street, Glasgow, UK G3 7HR; Institute for Social and Economic Research, University of Essex, Wivenhoe Park, Colchester, Essex, UK CO4 3SQ

**Keywords:** COVID-19, carers, mental health, inequalities, longitudinal

## Abstract

**Background:** Unpaid carers who look after another member of their household (home-carers) have poorer mental health than the general population. The first COVID-19 national lockdown led to an increasing reliance on home-carers and we investigate the short and longer-term impact of lockdown on their mental health.

**Methods:** Data from 9,737 adult participants (aged 16+) from the UK Household Longitudinal Study (Understanding Society) were used to explore changes in 12-item General Health Questionnaire (GHQ-12) score between (a) pre-pandemic (2019) and early lockdown (April 2020) and (b) early and later (July 2020) lockdown.

**Results:** GHQ-12 scores among home-carers were higher pre-lockdown and increased more than for non-carers from 2019 to April 2020 with further increases for home-carers compared with non-carers between April and July. Compared with respondents caring for a spouse/partner, those caring for a child under 18 had a particularly marked increase in GHQ-12 score between 2019 and April, as did those caring for someone with learning difficulties. Home-carers of children under 18 improved from April to July while those caring for adult children saw a marked worsening of their mental health. Home-carers with greater care burden saw larger increases in GHQ-12 score from 2019 to April and from April to July, and increases through both periods were greater for home-carers who had formal help prior to lockdown but then lost it.

**Conclusions:** The mental health of home-carers deteriorated more during lockdown than non-carers. Policies that reinstate support for them and their care-recipients will benefit the health of both vulnerable groups.

**What is already known on this topic:**

- Carers have poorer mental health than the general population.
- Among carers who live with the care recipient (home-carers), some subgroups have poorer mental health than others: female versus male; those who provide more hours of care and have been caring for longer; spousal carers compared with those caring for children (including adult), parents, or other relationships; those caring for individuals whose impairment results in behavioural disturbances, than those who care for individuals with physical or long-term health conditions.

**What this study adds:**

- In a large representative UK survey, the decline in mental health during lockdown was greater among home-carers than for the general population, and stayed poorer through to July, even as the general population’s mental health recovered slightly.
- Compared with respondents who were caring for a spouse/partner, those caring for a child under 18 had a particularly marked increase in GHQ-12 score between 2019 and April while those caring for an adult child experienced a substantial decline in their mental health between the beginning and end of the first lockdown (April to July).
- The increase in GHQ-12 in April from 2019 was highest among those caring for someone with a learning disability and lowest for those caring for someone with a problem related to old age.
- Home-carers who had a greater care burden, in terms of hours of care provided, or lost formal support during lockdown, had poorer mental health.

## Introduction

At the start of 2020 around 8.8 million adults in the UK were unpaid carers(Carers UK, 2019) supporting individuals, most commonly close family members, with disabilities, long-term health conditions, or needs related to old age. Around half were caring for someone living in the same household (home-carers) and half for someone living elsewhere. In spring 2020 the COVID-19 pandemic reached the UK and “lockdown” measures were introduced by the UK government. On 23rd March 2020, the Prime Minister announced the “Staying at home and away from others” policy, such that people could only leave home for limited purposes (shopping for basic necessities, one form of daily exercise, medical need, or to provide care or help for a vulnerable person).(https://www.gov.uk/government/speeches/pm-address-to-the-nation-on-coronavirus-23-march-2020; https://www.gov.uk/government/publications/full-guidance-on-staying-at-home-and-away-from-others/full-guidance-on-staying-at-home-and-away-from-others) This policy stayed in force for six weeks, then gradually relaxed to allow unlimited exercise, return to school for some pupils, non-essential shops re-opening, and people meeting outdoors. The major change in re-opening society came on 4th July, when the hospitality sector opened and two households could meet and stay overnight in same place.(https://www.health.org.uk/news-and-comment/charts-and-infographics/covid-19-policy-tracker) Alongside the lockdown restrictions, many non-COVID medical and social care services were withdrawn, cancelled, or changed from face-to-face to remote contact. These restrictions led to an increasing reliance by those with disabilities or ill health on informal support, with home-carers taking on a particularly important role. Results from a Carer’s UK survey in April 2020(Carers UK, 2020a) indicated that 70% of existing carers were providing more care during lockdown and more than a third were doing so as a result of changes to services. Moreover, a repeat survey in October 2020(Carers UK, 2020b) suggested that an additional 4.5 million new carers had been created by the pandemic.

Quarantine measures such as those imposed during the pandemic can have a negative psychological impact(Brooks et al., 2020) and concerns have been raised about the specific impact of COVID-19 lockdown measures on the mental health of the general population(Gunnell et al., 2020) as well as particular vulnerable groups including care recipients and their carers.(Holmes et al., 2020) Emerging evidence demonstrates that population mental health worsened in the early stages of lockdown(Niedzwiedz et al., 2020) although this may have reversed in the longer-term.(Chandola, Kumari, Booker & Benzeval, 2020) However, a number of carer-specific surveys have reported significant worsening of mental health both in the short(Carers UK, 2020a; Pavopoulou, Wood & Papadopoulos, 2020; Reaching Families, 2020) and longer term,(Carers UK, 2020b) although they do not make direct comparisons with the experiences of non-carers.

The current analysis builds on existing evidence to better understand the impact of COVID-19 and associated mitigation measures on the mental health of home-carers. Based on longitudinal data from Understanding Society annual interview and COVID-19 monthly web surveys, we focus on contemporaneous caring status during lockdown and compare the mental health of home-carers versus non-carers (or those only providing care outside the home) pre-pandemic and early (April) and later (July) in the first lockdown. We also acknowledge the wide range of experiences of home-carers and explore differences in mental health according to different carer characteristics (their relationship to the care recipient, the nature of the recipient’s health conditions and the caring burden and support received) to identify those at particularly high risk of poor mental health. We consider the following research questions: (i) how did the mental health of home-carers change during early and later lockdown relative to that of non-carers; (ii) how did the mental health of home-carers vary according to their different circumstances; and (iii) were differences among home-carers and between home-carers and non-carers explained by demographic differences?

## Methods

Data are from a substudy of the UK Household Longitudinal Study *(Understanding Society)* (UKHLS), a longitudinal, nationally representative study of the UK population.(Institute for Social and Economic Research, 2020a) All adults from households that took part in Waves 8 or 9 of the main survey were invited to take part in the COVID-19 April (24th-30^th^ inclusive) web survey (N= 42,330).(Institute for Social and Economic Research, 2020b) The COVID-19 survey has been repeated monthly and in July 2020 (24th-31^st^ inclusive), a module was added on caring responsibilities within the household. This paper uses data from the April and July COVID-19 surveys, and pre-pandemic data from 2019 (taken from wave 10 or 11 of the main survey based on the interview date).(University of Essex, 2020) Analyses are based on respondents who took part in all three.

Home-caring status in July was based on responses to the question: *Is there anyone living with you who is sick, disabled or elderly whom you look after or give special help to (for example, a sick, disabled or elderly relative, husband, wife or friend etc)*. Those giving a positive response were identified as home-carers and were asked how many people in the household they cared for and, if more than one, to focus on the main person they cared for. Additional information was collected on the care recipient’s condition (a long-term health condition (excluding mental health); a long-term mental health condition; a learning disability or developmental disorder such as autism; a physical disability; a problem related to old age; other condition); relationship to the home-carer (dependent child(ren) under 18; adult child(ren); parents or grandparents, including in-laws; siblings; spouse or partner; friends; other relatives; someone else); the hours per week spent caring; and any support they received with this from others in the household or from formal respite or support services such as day-care centres, school, college or carers supporting them in the home.

Respondents’ mental health at all three time points (2019, April 2020, July 2020) was measured using the 12-item General Health Questionnaire (GHQ-12), a widely used measure of non-psychotic psychological distress designed to capture depressive and anxiety symptoms.(Goldberg & Williams, 1988) Each item has four response categories on a Likert scale ranging from ‘not at all’ to ‘much more than usual’ (scored 0,0,1,1). Scores are then summed across items to produce a score between 0 and 12, with higher scores indicating poorer mental health. Primary analyses are based changes in scores so that changes in symptoms could be captured across the whole scale.

Analyses use ordinary least squares regression models comparing changes in GHQ-12 score between (a) pre-pandemic (2019) and early lockdown (April 2020) and (b) early and later (July 2020) lockdown, with all models adjusted for GHQ-12 scores at “baseline” ((a) 2019 and (b) April 2020). Preliminary models considered all respondents and compared changes in home-carers versus non-carers. Additional models were restricted to home-carers and compared changes according to their different circumstances. All models were then adjusted for age group (<40, 41-70, 71+) and sex to explore whether differences between home-carers and non-carers and between different groups of home-carers were driven by demographic differences.

Standard errors were adjusted to take account of the clustered and stratified sample and models included inverse probability weights to take account of unequal selection probabilities into the study and differential nonresponse at each wave, including to the COVID-19 survey. These weights ensure the results are reliable estimates representative of the UK adult population living in private households.(University of Essex, 2020) In supplementary analyses models were repeated based on GHQ-12 caseness with respondents scoring 4+ defined as having probable common mental disorder (CMD).

## Results

April web-survey interviews were completed (full and partial) by 17,761 participants and July web-survey interviews were completed (full and partial) by 13,754 participants, with 12,680 individuals taking part in both waves. Of these 12,209 (96%) participants also took part in a survey in 2019 (data from Waves 10-11). The current analyses are based on respondents with complete data available in 2019, April and July surveys. The weighted analytical sample is 9,737. However, in the weighted sample some participants had missing data on at least one covariate, and hence the weighted analytical sample for individual models varies between 9,369 and 9,614 depending on the covariates included.

Table 1 describes the basic characteristics of the analytic sample. The mean age of participants was 50.8 (standard deviation 15.8)) and 5,037 (52%) were female. In total 565 (6%) of respondents self-identified as home-carers in July 2020. Home-carers were older and more likely to be women than non-carers (22% versus 16% aged 71+ and 58% versus 52% female). Mean GHQ-12 scores were higher among home-carers versus non-carers in 2019 (2.45 versus 1.89). By April 2020 GHQ-12 scores in both groups had risen before falling again in July 2020. However, while July GHQ-12 scores among non-carers were similar to those in 2019, scores among home-carers continued to be higher than those measured prior to lockdown.

**Table 1.**
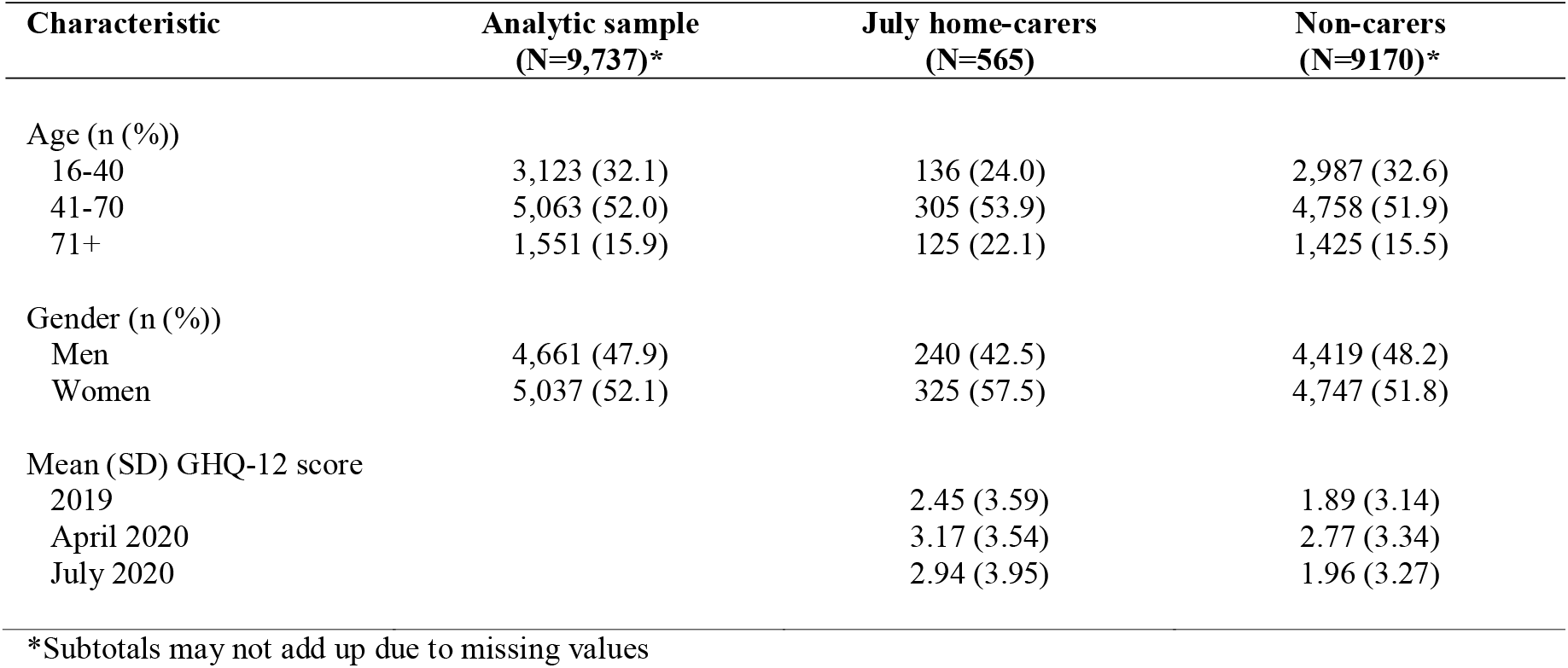
Characteristics of respondents who took part in 2019, April and July Covid-19 surveys.

Table 2 presents key characteristics of the 565 home-carers in July. Carers’ relationships with the care-recipient were: spouse/partner (41%), parent/grandparent (17%), child under 18 (15%), adult child (10%), and other (6%). Just over 10% of carers reported caring for more than one person. Care-recipients were reported as having a long-term health condition (45%), physical disability (31%), problems related to old age (23%), learning disability (23%), mental health condition (14%) or other (8%). The majority of home-carers (73%) reported also being a home-carer in 2019. Around 40% of home-carers provided care for under 20 hours a week with a further 38% providing 20-100 hours and 17% providing over 100 hours or continuous care. Most home-carers reported never receiving any state care/services before the pandemic (83%); however 10% stated that they received these services before the pandemic but lost it and 6% reported that they still received it. The majority did not share care responsibilities in the household (65%).

**Table 2.** Characteristics of home-carers who took part in 2019, April and July Covid-19 surveys.

| Characteristic | Analytical sample (home-carers in all three waves)<br>(N=565)* |
| --- | --- |
| Care recipient relationship |  |
| Child under 18 | 82 (14.6) |
| Adult child | 58 (10.3) |
| Parent or grandparent | 94 (16.8) |
| Spouse or partner | 232 (41.4) |
| Other | 33 ( 5.9) |
| Care for more than one person | 62 (11.1) |
| Care recipient condition** |  |
| Long term health condition | 257 (45.4) |
| Mental health condition | 79 (14.0) |
| Physical disability | 176 (31.2) |
| Learning disability | 127 (22.5) |
| Problems related to old age | 131 (23.2) |
| Other | 50 ( 8.0) |
| Carer in 2019 |  |
| Yes | 413 (73.1) |
| No | 152 (26.9) |
| Hours of care per week |  |
| Under 20 hours | 230 (40.8) |
| 20-100 hours | 213 (37.8) |
| Over 100 hours | 96 (17.0) |
| State care/services |  |
| Never had | 465 (83.3) |
| Had before lockdown and lost | 58 (10.4) |
| Had before lockdown and still do | 32 ( 5.7) |
| Share care responsibilities |  |
| Yes | 196 (34.8) |
| No | 368 (65.2) |

\*Subtotals may not add up due to missing values; \*\*Total is more than 100 as question is "please select all that apply"

Table 3 presents changes in GHQ-12 scores from (a) 2019 to April and (b) April to July in home-carers versus non-carers. Mean scores among home-carers increased slightly more than for non-carers from 2019 to April and this was marginally more marked after adjustment for age and sex (change in GHQ-12 score in home-carers versus non-carers: 0.32 (0.04, 0.68)). However, increases from April to July were greater among home-carers when compared with non-carers (0.80 (0.34, 1.25)).

**Table 3.** Difference in GHQ-12 score from (a) baseline to April and (b) April to July according to home-carer status.

| Caring status in July | Baseline to April |  | April to July |  |
| --- | --- | --- | --- | --- |
|  | Adjusted for baseline<br>GHQ-12 | Additionally adjusted<br>for age and sex | Adjusted for April<br>GHQ-12 | Additionally adjusted<br>for age and sex |
| Non-carer | 0.00 | 0.00 | 0.00 | 0.00 |
| Home carer | 0.26 (-0.09, 0.61) | 0.32 (0.04, 0.68) | 0.77 (0.32, 1.23) | 0.80 (0.34, 1.25) |

Table 4 focuses on home-carers and examines changes in GHQ-12 score according to their different characteristics. There was little evidence of a difference in GHQ-12 scores by home-carers’ age. However, home-carers who were women saw a greater increase in GHQ-12 score between 2019 and April 2020 when compared to men (1.13 (0.45, 1.81)); this difference by sex was not evident in analyses of change in GHQ-12 score between April and July.

**Table 4.** Difference in GHQ-12 score from (a) baseline to April and (b) April to July according to home-carer characteristics.

| Caring status in July | Baseline to April |  | April to July |  |
| --- | --- | --- | --- | --- |
|  | Adjusted for baseline GHQ-12 | Additionally adjusted for age and sex | Adjusted for April GHQ-12 | Additionally adjusted for age and sex |
| Age group |  |  |  |  |
| <40 | 0.00 | - | 0.00 | - |
| 41-70 | 0.40 (-0.45, 1.24) |  | 0.18 (-0.85, 1.20) |  |
| 71+ | -0.53 (-1.39, 0.33) |  | -0.36 (-1.06, 0.34) |  |
| Sex |  |  |  |  |
| Male | 0.00 | - | 0.00 | - |
| Female | 1.13 (0.45, 1.81) |  | 0.38 (-0.55, 1.30) |  |
| Relationship of care recipient to carer |  |  |  |  |
| Spouse/partner | 0.00 | 0.00 | 0.00 | 0.00 |
| Child under 18 | 0.91 (0.30, 1.52) | 0.77 (0.10, 1.44) | -0.28 (-0.44, 0.12) | -0.43 (-0.65, -0.20) |
| Adult child | 0.39 (0.12, 0.66) | -0.14 (-0.40, 0.11) | 2.75 (2.42, 3.08) | 2.64 (2.30, 2.99) |
| Parent/grandparent | 0.01 (-0.35, 0.37) | -0.18 (-0.58, 0.22) | 0.58 (0.26, 0.90) | 0.44 (0.09, 0.79) |
| Other | -0.57 (-0.76, -0.37) | -0.71 (-0.93, -0.49) | -0.16 (-0.32, 0.00) | -0.26 (-0.45, -0.07) |
| Care for more than one person | 1.54 (1.23, 1.86) | 1.38 (0.96, 1.81) | 1.21 (0.77, 1.65) | 1.04 (0.55, 1.53) |
| Care recipient has long term health condition |  |  |  |  |
| No | 0.00 | 0.00 | 0.00 | 0.00 |
| Yes | -0.14 (-0.44, -0.17) | 0.01 (-0.31, 0.32) | 0.57 (0.37, 0.78) | 0.63 (0.42, 0.83) |
| Care recipient has mental health condition |  |  |  |  |
| No | 0.00 | 0.00 | 0.00 | 0.00 |
| Yes | 0.35 (-0.02, 0.72) | 0.22 (-0.13, 0.56) | 0.57 (0.29, 0.85) | 0.57 (0.27, 0.87) |
| Care recipient has learning disability |  |  |  |  |
| No | 0.00 | 0.00 | 0.00 | 0.00 |
| Yes | 1.06 (0.66, 1.46) | 0.91 (0.53, 1.29) | 1.32 (1.03, 1.60) | 1.27 (0.99, 1.56) |
| Care recipient has physical disability |  |  |  |  |
| No | 0.00 | 0.00 | 0.00 | 0.00 |
| Yes | -0.10 (-0.44, 0.24) | -0.08 (-0.39, 0.24) | 0.70 (0.51, 0.88) | 0.70 (0.51, 0.89) |
| Care recipient has problem related to old age |  |  |  |  |
| No | 0.00 | 0.00 | 0.00 | 0.00 |
| Yes | -0.56 (-0.81, -0.31) | -0.46 (-0.71, -0.21) | -0.66 (-0.93, -0.38) | -0.61 (-0.92, -0.30) |
| Care recipient has other condition |  |  |  |  |
| No | 0.00 | 0.00 | 0.00 | 0.00 |
| Yes | 0.03 (-0.79, 0.85) | -0.17 (-0.86, 0.52) | -0.11 (-0.76, 0.55) | -0.19 (-0.83, 0.46) |
| Carer in 2019 |  |  |  |  |
| No | 0.00 | 0.00 | 0.00 | 0.00 |
| Yes | -0.01 (-0.62, 0.60) | -0.07 (-0.53, 0.67) | -0.24 (-0.99, 0.50) | -0.21 (-0.97, 0.55) |
| Hours of caring per week |  |  |  |  |
| <20 | 0.00 | 0.00 | 0.00 | 0.00 |
| 20-100 | 0.57 (0.31, 0.83) | 0.29 (0.03, 0.55) | 0.49 (0.27, 0.71) | 0.42 (0.19, 0.66) |
| 101+ | 0.67 (0.13, 1.20) | 0.46 (-0.04, 0.96) | 1.09 (0.77, 1.41) | 1.10 (0.76, 1.43) |
| State care/services |  |  |  |  |
| Never had | 0.00 | 0.00 | 0.00 | 0.00 |
| Had and lost | 0.70 (0.27, 1.14) | 0.54 (0.13, 0.95) | 2.04 (1.51, 2.56) | 1.98 (1.46, 2.49) |
| Still have | 0.09 (-1.44, 1.45) | 0.07 (-1.29, 1.43) | -0.22 (-0.56, 0.13) | -0.20 (-0.54, 0.15) |
| Share caring responsibilities |  |  |  |  |
| No | 0.00 | 0.00 | 0.00 | 0.00 |
| Yes | 0.34 (0.10, 0.58) | 0.49 (0.26, 0.72) | 0.07 (-0.16, 0.30) | 0.08 (-0.15, 0.32) |

Compared with respondents who were caring for a spouse/partner, those caring for a child under 18 had a particularly marked increase in GHQ-12 score between 2019 and April (0.91 (0.30, 1.52)) with those caring for “other” household members having a less marked increase. These differences were much reduced in comparison of GHQ-12 scores from April to July; however, home-carers responsible for adult children had considerably greater increases in GHQ-12 score over this period (2.75 (2.42, 3.08)). Home-carers who cared for more than one person in the household had markedly higher increases in GHQ-12 scores in April (1.54 (1.23, 1.86)) and the same was true comparing July with April scores (1.21 (0.77, 1.65)). The increase in GHQ-12 in April from 2019 was highest among those caring for someone with a learning disability (1.06 (0.66, 1.46)) and lowest for those caring for someone with a problem related to old age (−0.56 (−0.81, -0.31)). Looking at change in GHQ-12 from April to July, those caring for someone with a learning disability continued to have the greatest increase (1.32 (1.03, 1.60)). However, increases were also marked for those caring for individuals with long term (0.57, 0.37, 0.78)) and mental health conditions (0.57 (0.29, 0.85)) and physical disabilities (0.70 (0.51, 0.88)), while those caring for someone with a problem related to old age saw a relative decline in score (−0.66 (−0.93,-0.38)). There was very little difference in the changes in GHQ-12 scores in either period comparing existing and new home-carers. Home-carers who provided higher numbers of hours of care per week saw a corresponding increase in their GHQ-12 scores from 2019 to April and, more markedly, from April to July (e.g. relative to those providing <20 hours, change from April to July: 0.49 (0.27, 0.71) and 1.09 (0.77, 1.41) for those providing 20-100 and 100+ hours respectively). Increases in GHQ through both periods were greater for home-carers who had had help from state care and services prior to lockdown but then lost it (e.g. April to July change relative to those who had never had support: 2.04 (1.51, 2.56)). Home-carers who shared responsibilities with others in the household had a somewhat greater increase in GHQ-12 from 2019 to April but this difference was no longer evident when comparing April to July. Adjusting for age and sex had very little effect on the differences between groups.

Results from sensitivity analyses using 4+ symptoms as indicative of probable common mental disorder (CMD) were similar to those reported above (Appendix Tables X1 and X2). However, unadjusted odds ratios suggest that, compared with spousal carers, carers of adult children in April and carers of parents or grandparents in July were 2.00 (1.64, 2.47) and 1.59 (1.01, 2.49) times more likely respectively to show signs of psychological distress. Additionally, in July, those caring for someone with a mental health of long-term condition were 1.70 (1.25, 2.33) and 1.56 (1.27, 1.91), times more likely respectively to show signs of psychological distress than those caring for someone without these conditions.

## Discussion

Carers and care recipients have been identified as vulnerable groups in terms of the mental health consequences of COVID-19 and associated mitigation measures(Holmes et al., 2020) with an urgent call for research to better understand their experiences. We found that home-carers had worse mental health than non-carers prior to the pandemic. By April 2020 mental health had deteriorated in both groups but to a greater extent among home-carers and while there was some improvement in mental health between April and July 2020 in both groups this was more modest amongst home-carers. Home-carers were older and more likely to be women than non-carers but differences in GHQ scores were not explained by these factors.

Among home-carers in the current analyses, some were particularly badly affected by lockdown. Early declines in mental health in April were more marked in female carers, although these differences were no longer evident by July. Perhaps unsurprisingly, those caring for more than one person and for more hours had particularly marked declines in mental health in April compared with pre-pandemic levels and further declines by July. In addition, those who had lost respite care also fared worse in April and had a further marked worsening of mental health by July. Around a quarter of those self-identifying as a home-carers in July had not identified as a home-carer prior to the pandemic but there was no evidence to suggest that this group fared better or worse than pre-existing carers. In general, these findings are consistent with reported differences between carers pre-pandemic. Previous studies have showed that mental health was worse among carers who live with the care recipient,(Shah, Wadoo & Latoo, 2010) female versus male carers,(Shah et al., 2010; Schulz & Sherwood, 2008; Pinquart & Sorensen, 2003) and for carers who provide more hours of care and have been caring for longer.(Schulz & Sherwood, 2008)

During the first lockdown, our study shows parents of children under 18 had markedly greater declines in mental health in April 2020 when compared with spousal or other home-carers but by July this same group had seen an improvement compared with others. This contrasts with findings pre-pandemic, with spousal carers generally reported as having worse mental health than those caring for children (including adult), parents, or other relationships.(Schulz & Sherwood, 2008; Pinquart & Sorensen, 2003) This difference may reflect the significant impact of school closures and subsequent partial reopening on parent carers. In contrast, the mental health of parents caring for adult children was not markedly worse than other home-carers in April but saw a substantial decline by July, suggesting that their negative experiences were continuing and, indeed, worsening as lockdown progressed. This is consistent with the continuing closure of day and respite services, in contrast to schools re-opening, and reports of increases in challenging behaviours in those with autism and learning difficulties, as well as ongoing difficulties in accessing food and other supplies.(Carers UK, 2020a; Reaching Families, 2020)

Existing evidence also suggests that those who care for individuals whose impairment results in behavioural disturbances, for example mental health problems, dementia or cognitive impairment, have worse mental health than those who care for individuals with physical or long-term health conditions.(Shah et al., 2010; Schulz & Sherwood, 2008; Pinquart & Sorensen, 2003) In the current analyses, respondents caring for individuals with learning disabilities had the greatest declines in mental health early in lockdown. The further exacerbation of their mental health may again reflect the sudden closure of schools and day services; the withdrawal of respite care has been highlighted as a particular issue by those caring for individuals with learning difficulties or autism(Pavlopoulou et al., 2020; Reaching Families, 2020) whereas health services for those with physical health problems were more likely to continue, albeit in a reduced or limited form. In addition, carers for those with learning difficulties reported additional challenges as a result of their care-recipient not understanding new, often restricted, circumstances or guidance on hygiene and social distancing.(Carers UK, 2020a)

By July home-carers for all conditions other than those associated with old age had seen a marked decline in their mental health, highlighting the ongoing cumulative strain of caring under lockdown conditions arising from, for example, anxiety regarding infection among those caring for someone who was shielding,(Office for National Statistics, 2020) or problems accessing non-COVID health care. It has been suggested that individuals with pre-existing mental health problems might have been particularly vulnerable to COVID-19 mitigation measures(Brooks et al., 2020; Gunnell et al., 2020; Holmes et al., 2020) and disruptions to mental health services, particularly those based in the community,(World Health Organisation, 2020) and our results suggest that home-carers for those with mental health problems had particularly poor mental health themselves by July.

The current analyses are based on a large longitudinal dataset including specific detailed questions regarding respondents’ caring experiences during lockdown and use of a validated mental health measure in repeated waves before and throughout the lockdown period. However, there are also some limitations. Comparison with caring status in pre-pandemic waves suggests that home-carers may have been under-represented in the COVID surveys; however, the use of analytical weights means that our results are likely to be generalizable to the UK population. In contrast to previous work,(Gallagher & Wetherell, 2020) we considered caring status during rather than before lockdown as the pandemic created new caring roles.(Carers UK, 2020b) However, this was measured in July, and so may not accurately reflect circumstances in April although the error is likely to be small and results for new and pre-existing home-carers were almost identical. Although detailed data were available on the caring experiences of respondents during the pandemic these were not exhaustive; for example we lacked information regarding the type of care provided. Finally, we consider mental health at the beginning of the first lockdown and as it progressed. However, given further national lockdowns, it will be important to revisit these issues to investigate the short- and longer-term impacts of repeated restrictions and reduced support on the mental health of carers.

## Conclusion

Informal home-carers have been described as “the forgotten health-care workers during the COVID-19 pandemic”(Chan et al., 2020) Our results clearly demonstrate that the mental health of home-carers have been disproportionately affected by COVID-19 and associated mitigation measures. Responses to the pandemic have largely focussed on infection control but there is also growing recognition of the need to support mental health. Our study shows this is particularly an issue for those caring for others in the home and, as the pandemic and mitigations continue, better policies that support mental health will be required. The restarting and maintenance of relevant health and social care services should be a priority to reduce the burden on informal home-carers, with consideration given to how to support carers of those with conditions such as dementia and learning disabilities, whose mental health seems to be particularly affected.

There are millions of unpaid home-carers in the UK who provide vital support to members of their household. Although there is good existing evidence that this impacts negatively on their mental health, policies and services to support these individuals are limited. Our findings, along with those from other COVID-specific research, shine a spotlight on the challenges faced by this neglected group. The post-COVID restart of health and social services provides an ideal opportunity to recognise the role of home-carers and to consider how best to support them.

## Data Availability

The research data are distributed by the UK Data Service.

## Contributors

All authors designed the study and contributed equally to this paper, and collectively drafted paper. KSR implemented the statistical analyses. All authors critically revised the article, contributed to data interpretation, and finalised and approved the manuscript.

## Ethical approval

Ethics approval was granted by the University of Essex Ethics Committee for the COVID-19 surveys (ETH1920-1271).

### Appendix tables

**Table X1:**
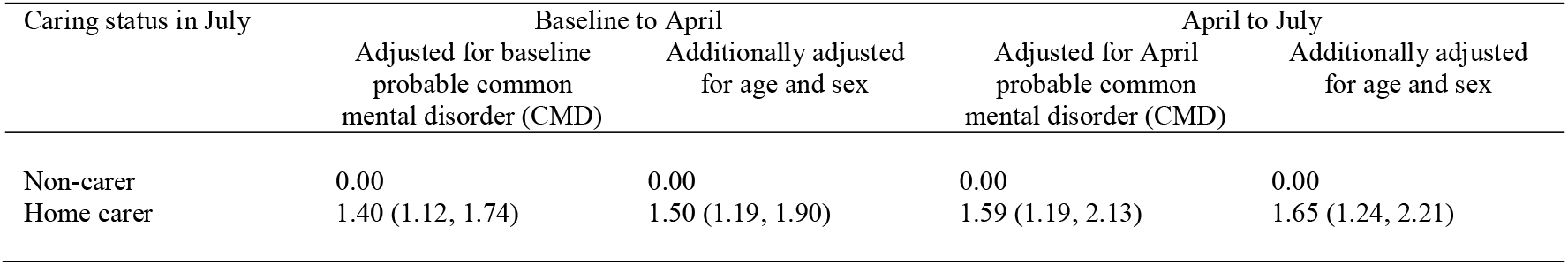
Odds of probable common mental disorder (CMD) score from (a) baseline to April and (b) April to July according to home-carer status.

| Caring status in July | Baseline to April |  | April to July |  |
| --- | --- | --- | --- | --- |
|  | Adjusted for baseline probable common mental disorder (CMD) | Additionally adjusted for age and sex | Adjusted for April probable common mental disorder (CMD) | Additionally adjusted for age and sex |
| Non-carer | 0.00 | 0.00 | 0.00 | 0.00 |
| Home carer | 1.40 (1.12, 1.74) | 1.50 (1.19, 1.90) | 1.59 (1.19, 2.13) | 1.65 (1.24, 2.21) |

**Table X2:**
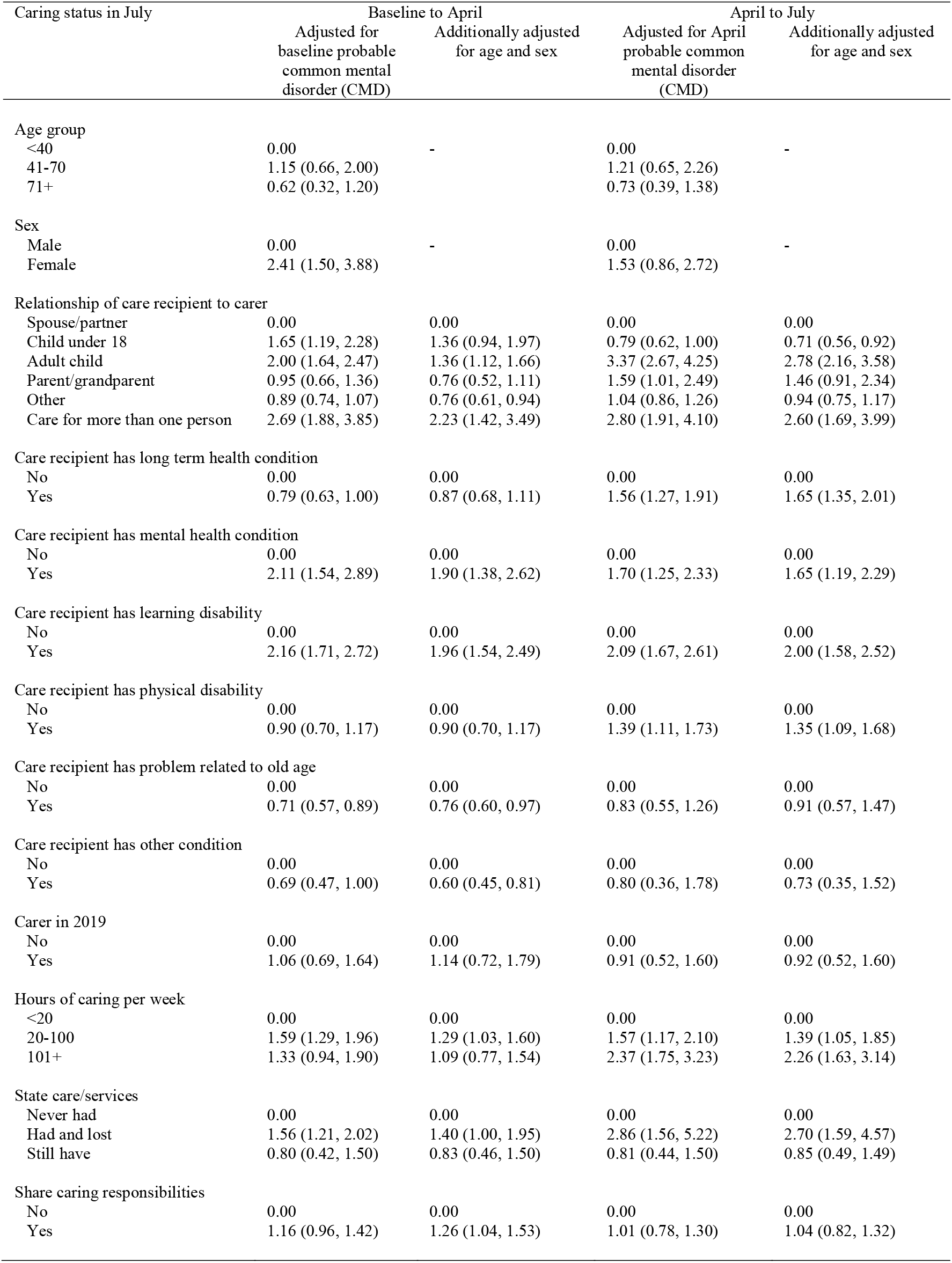
Odds of probable common mental disorder (CMD) score from (a) baseline to April and (b) April to July according to home-carer characteristics.

## References

Brooks SK, Webster RK, Smith LE, Woodland L, Wessely S, Greenberg N, … Rubin G. (2020) The psychological impact of quarantine and how to reduce it: rapid review of the evidence. Lancet, 395(10227), 912–20.

Carers UK. (2019) Facts about carers. London, UK: Carers UK.

Carers UK. (2020a) Caring behind closed doors. Forgotten families in the coronavirus outbreak. London, UK: Carers UK.

Carers UK. (2020b) Caring behind closed doors: six months on. London, UK: Carers UK.

Chan EYY, Gobat N, Kim JH, Newnham EA, Huang Z, Hung H, … Wong SYS. (2020) Informal home care providers: the forgotten health-care workers during the COVID-19 pandemic. Lancet, 2020, 395(10242), 1957–9.

Chandola T, Kumari M, Booker CL, Benzeval M. (2020) The mental health impact of COVID-19 and lockdown-related stressors among adults in the UK. Psychol Med, 1–10.

Gallagher S, Wetherell MA. (2020) Risk of depression in family caregivers: unintended consequence of COVID-19. B J Psych Open, 6(6), e119.

Goldberg D, Williams D. (1988) User’s guide to the General Health Questionnaire. 1988. London, UK: University of London, Institute of Psychiatry.

Gunnell D, Appleby L, Arensman E, Hawton K, John A, Kapur N, … Pirkis J (2020). Suicide risk and prevention during the COVID-19 pandemic. Lancet Psychiatry, 7(6), 468–71.

Holmes EA, O’Connor RC, Perry VH, Tracey I, Wessely S, Arseneault L, … Bullmore E (2020) Multidisciplinary research priorities for the COVID-19 pandemic: a call for action for mental health science. Lancet Psychiatry, 7(6), 547–60. https://www.gov.uk/government/publications/full-guidance-on-staying-at-home-and-away-from-others/full-guidance-on-staying-at-home-and-away-from-others

https://www.gov.uk/government/speeches/pm-address-to-the-nation-on-coronavirus-23-march-2020

https://www.health.org.uk/news-and-comment/charts-and-infographics/covid-19-policy-tracker

Institute for Social and Economic Research. (2020a) Understanding Society: Waves 1-10, 2009-2019 and Harmonised BHPS: Waves 1-18, 1991-2009, User Guide, 29 October 2020. Colchester, UK: University of Essex.

Institute for Social and Economic Research. (2020b) Understanding Society COVID-19 User Guide. Version 5.1. December 2020. Colchester, UK: University of Essex.

Niedzwiedz CL, Green MJ, Benzeval M, Campbell D, Craig P, Demou E, … Katikireddi SV (2020) Mental health and health behaviours before and during the initial phase of the COVID-19 lockdown: longitudinal analyses of the UK Household Longitudinal Study. J Epidemiol Community Health. doi: 10.1136/jech-2020-215060

Office for National Statistics. (2020) Coronavirus and the social impacts on disabled people in Great Britain. April 2020. London, UK: Office for National Statistics.

Pavlopoulou G, Wood R, Papadopoulos C. (2020) Impact of Covid-19 on the experiences of parents and family carers of autistic children and young people in the UK. London, UK: UCL Institute of Education.

Pinquart M, Sorensen S. (2003) Differences between caregivers and noncaregivers in psychological health and physical health: a meta-analysis. Psychol Aging, 18(2), 250–67.

Reaching Families. (2020) Coronavirus Survey Report. Empowering families of children and young people with special educational needs and disabilities in West Sussex. Worthing, UK: Reaching Families.

Schulz R, Sherwood PR. (2008) Physical and mental health effects of family caregiving. Am J Nurs, 108(9 Suppl), 23–7.

Shah AJ, Wadoo O, Latoo J. (2010) Psychological distress in carers of people with mental disorders. British Journal of Medical Practitioners, 3(3), 327.

University of Essex, Institute for Social and Economic Research. (2020) Understanding Society: COVID-19 Study, 2020. [data collection]. 6th Edition. UK Data Service. SN: 8644, 10.5255/UKDALJSNLJ8644LJ6.

World Health Organisation. (2020) The impact of COVID-19 on mental, neurological and substance use services: results of a rapid assessment. Geneva: World Health Organisation.

